# Treating malignant hypertension with a low-sodium, low-protein, low-fat diet

**DOI:** 10.1101/2025.04.01.25325063

**Authors:** Scott L. Sanoff, Philip J. Klemmer, Francis A. Neelon, Jong Ok La, David Lopez, Anastacia Bohannon, William McDowell, Fredrich C. Luft, Yi-Ju Li, Lin Pao-Hwa

**Author notes:** Corresponding Authors: Pao-Hwa Lin, PhD, DUMC 3487, Durham, NC 27710.

## Abstract

**Background:** The Rice Diet (RD), a low-sodium (<150mg/day), low-protein (20g/day), low-fat (<5g/day), diet was used to treat patients with malignant hypertension (MH) beginning in the 1940’s, before any effective anti-hypertensive drugs were available. We retrospectively analyzed a curated cohort of RD patients with MH to assess factors, including dietary adherence, associated with blood pressure (BP) reduction.

**Methods:** From 17,487 RD charts, we identified 544 MH patients (baseline systolic BP (SBP) ≥170 mmHg and with concurrent retinal hemorrhage and/or papilledema), excluding those with diabetes, brain tumor, or prior sympathectomy. Outcome data were censored after any 30-day break in consecutive data. Baseline features, BP changes from baseline to Week 4, and adherence (assessed by urinary chloride, UCl) were evaluated using summary statistics, univariate and multivariable analyses.

**Results:** Most patients participated in the RD program before antihypertensive drugs were available; only 48 (4.2%) received any anti-hypertensive medications in the first month. The cohort (68.9% male) had a median baseline BP of 213/128 mmHg and BMI of 23.6 kg/m^2^. Median time in the program before censoring was 109 days; median total amount of time in the RD program was 333 days. BP declined significantly within the first week, reaching 179/108 mmHg at Week 4. UCl dropped from 217 to 21 mg/dL by Week 4. Lower UCl, higher baseline BP and female gender, but not retinal hemorrhage and/or papilledema, were associated with greater SBP reduction.

**Conclusion:** The low-sodium, low-fat, low-protein RD effectively lowered BP in patients with MH in four weeks, independent of antihypertensive medications.

## Background

The term ‘malignant hypertension’ (MH) was introduced into the English-language literature by Drs. Wagener and Keith in 1924 to describe patients with high blood pressure (BP), diffuse vascular injury, retinal hemorrhage with or without papilledema, rapid clinical decline,^1^ and a poor prognosis. In 1928 Keith, Wagener and Kernohan reported that in a cohort of 81 patients, only “five patients lived two years or longer.”^2^ Prior to the approval of chlorothiazide by the Food and Drug Administration (FDA) in 1958,^3^ persons with MH had no proven therapeutic options.

The Rice Diet (RD), was devised in the early 1940’s by Dr. Walter Kempner to treat patients with uncontrolled hypertension.^4^ Following early reports of its success,^5^ thousands of persons from the United States and other countries came to Durham, NC, to participate in the RD program. The clinical records of those patients provide a unique opportunity to study the impact of this restrictive diet without the confounding effect introduced by the concomitant use of modern anti-hypertensives.^6-8^ Despite a drastic decrease in the prevalence of MH over the past decades, MH is not extinct and may be increasing in countries with less advanced health care resources.^9,10^

Since 1950, nearly 200 randomized trials have examined the biological impact of low sodium diets. To the best of our knowledge, and despite there being no established lower limit to the BP lowering effects of sodium restriction,^11^ none have evaluated a diet as restricted in sodium as the RD. Furthermore, each of these studies has had to calibrate its equipoise to accommodate use of anti-hypertensive medications, thereby obscuring the effects of diet per se on patients with MH.^11,12^ In addition, although case series have reported outcomes of RD program participants,^6^ no publication has described the impact of the RD on the full cohort of enrolled patients with MH.

We digitized the treatment records of 17,487 RD program participants, and from these records, have constructed a database capturing key patient-level data elements.^13^ Here we present a retrospective interventional cohort study examining the BP response of all RD program participants with MH, the relationship between participants’ estimated sodium intake and BP responses, and patient factors that predict BP lowering.

## Methods

### Study Design

The database was derived from patient-level treatment records maintained by Dr. Kempner and the clinical staff of the RD program. This study focuses on patients with MH enrolled in the RD program regarding (1) weekly systolic (SBP) and diastolic (DBP) blood pressure trends over four weeks following diet initiation; (2) the relationship between SBP and sodium intake (using urinary chloride [UCl] as a marker of sodium intake, because of an ability to measure urine sodium directly) over the first four weeks of RD treatment; (3) the association between patient-level factors and BP changes from entry to week 4, and (4) the above associations by MH classes (see Data analysis). Construction of this database and the execution of this study were approved by the Duke University Medical Center IRB (Pro00105257).

### Study Intervention

The RD was conceived by Dr. Kempner based on his studies of in vitro renal cellular metabolism. This restricted diet contained <5g/day of fat, approximately 20 gm/day of protein, <150 mg/day of sodium, and <200 mg/day of chloride. Nominal daily energy intake of approximately 2,000 Kcal was tailored to meet individual patient weight targets.^4^ Fluid intake was limited to 0.7-1 L/day of fruit and fruit juices in order to avoid water intoxication from the low solute content of the diet. The diet was supplemented with iron, vitamins A and D, thiamine, riboflavin, and niacinamide. The RD comprised approximately 250-350 grams of dry rice daily, boiled or steamed in water or fruit juice. All fruit juices and fruits except tomato and vegetable juices, nuts, dates, avocados, and canned or dried fruit or fruit derivatives containing anything other than white sugar or dextrose were allowed. Patients’ clinical status and various biochemical variables were monitored closely during initiation of the diet, often while in the hospital. If response to the diet proved to be adequate and durable (over months), small amounts of non-leguminous vegetables, potatoes, lean meat or fish were sometimes added.^7^

### Study Population

All adult patients (age ≥18yo) enrolled in the RD program were considered for this report. Those with at least one SBP ≥170mmHg between day -7 and day 6 of the RD start date, *and* the presence of retinal hemorrhage and/or papilledema on funduscopic examination between day -30 and day 30 of starting the RD were considered for analysis. Photographic retinal images were obtained and analyzed by a dedicated ophthalmologist. Patient data were censored if there was a data entry gap of >30 days, and only data collected before the 30-day gap were analyzed. Each patient was considered only once, and only the first visit to the RDP was analyzed for patients who returned after a 30-day break. To minimize misclassification and confounding, we excluded: patients with hyperglycemia (to avoid including diabetic retinopathy), brain tumor, or previous sympathectomy.

### Statistical Analysis

Median (and Interquartile Range; IQR) for continuous variables, and frequency (percentage) for categorical variables were used to describe the cohort demographics. Patients with MH were further classified as described by Keith, Wagener, Barker (KWB) having Class III hypertension (with retinal hemorrhage only), Class IV (with both retinal hemorrhage and papilledema), or what we have designated Class V (papilledema without hemorrhage), a class not described by KWB.^14^

Baseline SBP, DBP, urine concentration of chloride (UCl), and blood concentration of non-protein nitrogen (NPN; laboratory reference range 25-35 mg/dL) were defined as the average of available data recorded between day -7 to day +1. Baseline weight is the average of weights from day -2 to day +1. The trends of SBP and DBP from baseline to Week 4 are graphically shown as the weekly average (e.g. Week 1 BP is the average of BPs recorded between day 1 and day 7) with 95% confidence intervals (CI). Median (IQR) of SBP, DBP, and UCl were also computed for each weekly timepoint from baseline to Week 4.

We further defined adherence to RD by the number of weeks each participant met the goal of UCL ≤42 mg/dL, an optimal cut-point determined from UCl changes and associated SBP responses during the first 4 weeks. Participants were classified into three adherence groups: High adherence (UCl goal was met for at least 3 of the 4 weeks); moderate adherence (goal was met for 1 or 2 weeks); and low adherence (goal was never achieved during the 4 weeks). Dietary adherence was also calculated as the percent of available UCl values ≤42 mg/dL for patients with at least 6 UCl values available between week 1 and week 4. Univariable linear mixed model was used to regress weekly SBP changes from the baseline on their corresponding time of measurement (i.e., week) with random intercept by patient to assess rate of SBP change for each adherence group.

Univariable linear regression was used to identify the relationship between baseline characteristics (age, gender, retinal hemorrhage, papilledema, SBP, NPN, UCl, dietary adherence and weight) and SBP changes from baseline to week 4 (or week 3 if no week 4 data were available). Baseline characteristics with p-value less than 0.15 in the univariable analysis were used for multivariable linear regression modeling, where variables with p < 0.05 were considered significant. To evaluate any possible influence of BP medications, sensitivity testing was conducted in patients who were not taking BP medications during the first 4 weeks.

Lastly, we defined responders to the RD as those who achieved a 5% or greater BP reduction from baseline to week 4 and compared their characteristics to those of non-responders (<5% BP reduction).

## Results

### Demographics and Baseline Values

The RD program database contains data from 16,266 adults (**Figure 1**). Of the 807 patients with SBP ≥ 170 mmHg between day -7 and +6 and with concomitant retinal hemorrhage, papilledema, or both, 232 were excluded because of one or more of the following: overt diabetes or hyperglycemia (N=228), prior sympathectomy (N=6), brain tumor (N=2); another 31 were excluded because they participated in the RD program for <7 days. The final cohort of 544 analyzable patients with MH includes 311 with retinal hemorrhage (KWB Class III), 211 with hemorrhage and papilledema (KWB Class IV), and 22 with papilledema only (Class V).

**Figure 1.**
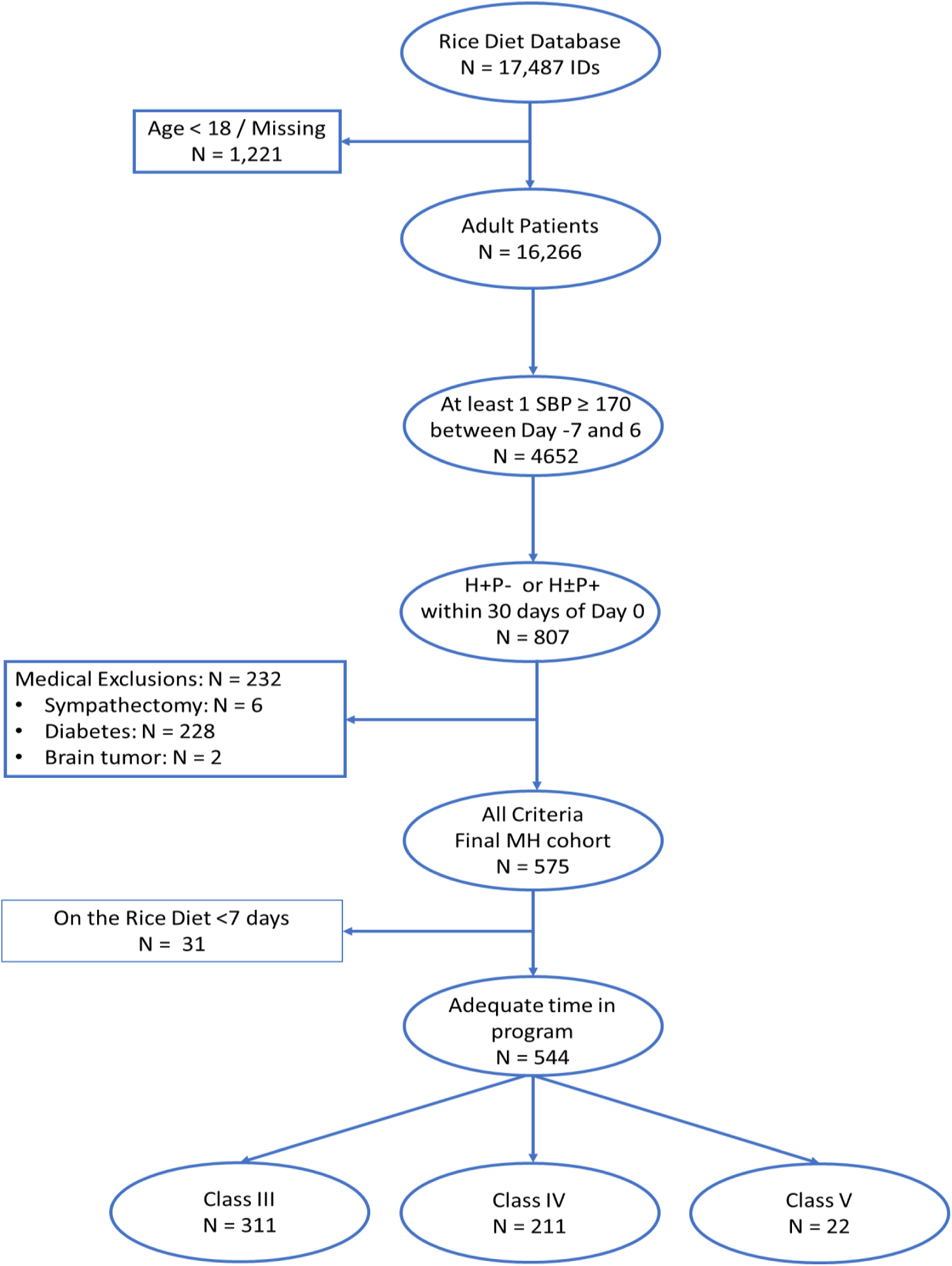
CONSORT Diagram.

**Figure 2** shows the number of patients with MH who attended the RD program for the first time per 3-year interval. Few participated during 1940-1942, but the number increased thereafter, peaking during 1949-1951. Subsequently, patient arrivals dropped drastically until around 1955, after which the rate of accrual dropped slowly but did not entirely disappear.

**Figure 2.**
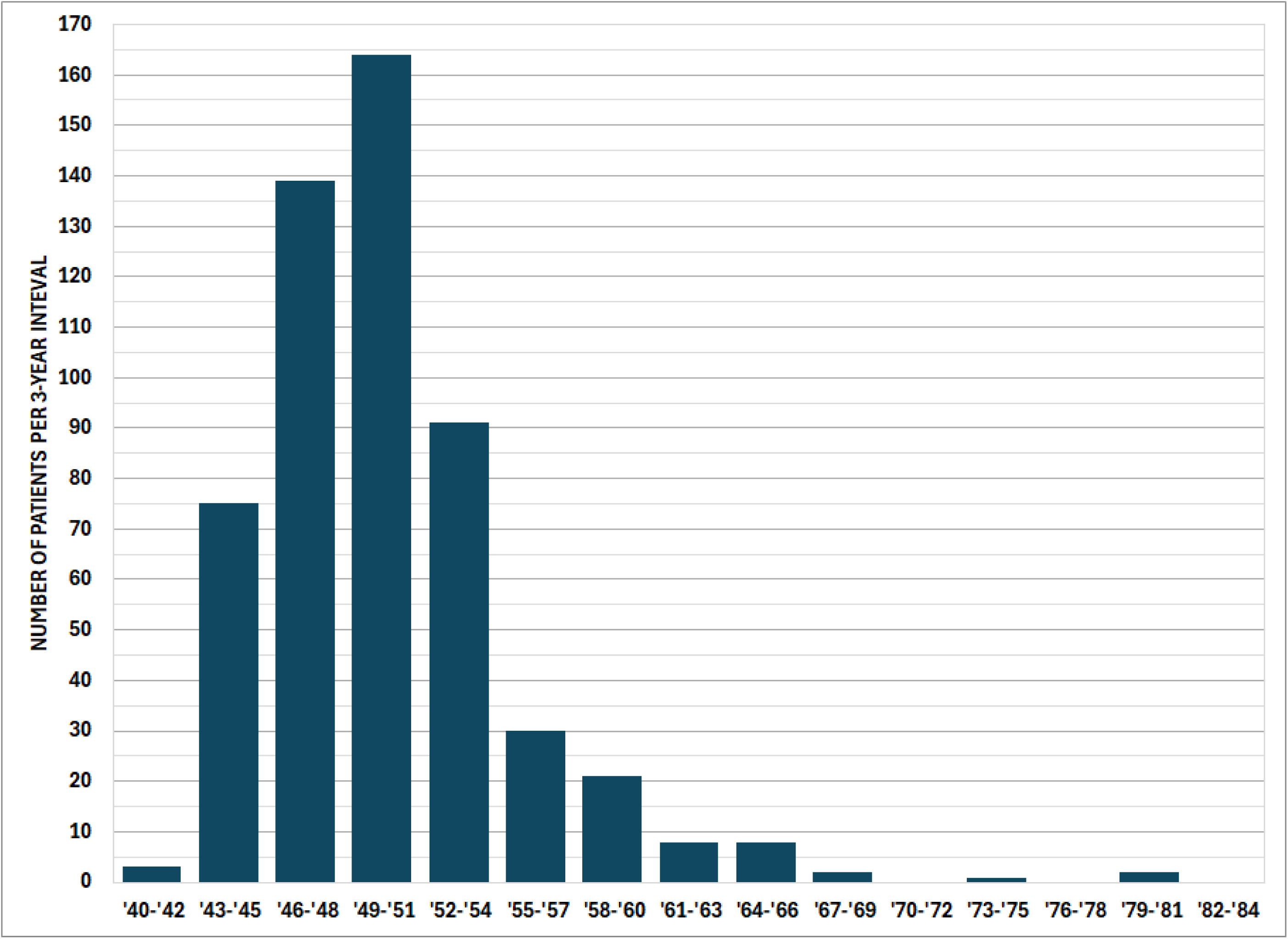
Number of participants first attending the Rice Diet Program per 3-year interval.

Demographic and clinical characteristics for the cohort are displayed in **Table 1**. Patients were predominately male (68.9%); median age was 50 (range: 19, 81), and more than 90% of the cohort was enrolled before effective anti-hypertensive medication was introduced in 1958. The median (IQR) for key variables is as follows: days spent in the program before a 30-day or longer data break was 109 days (46.0, 176); baseline SBP was 213 mmHg (194, 232) and DBP, 128mmHg (114, 140); baseline weight 69.2 kg (60.2, 78.1) and BMI 23.6 (21.5, 27.3) kg/m^2^ (unfortunately, a large number of patients had no recorded weight [61.8 %] or height [23.5%] data); baseline NPN was 41.0 mg/dL (35, 57) and 24 hour UCl was, 217 mg/dL (113, 356). Presuming patients were at ‘steady state’ and making approximately 1 liter of urine per day, this UCl is consistent with a sodium intake of approximately 1,430 mg per day at study entry. We believe that this value may be low due to sodium restriction in some patients prior to starting the RD.

**Table 1.** Rice Diet Malignant Hypertension Cohort Baseline Characteristics.

|  | Class III (H+P-)<br>N = 311 | Class IV (H+P+)<br>N = 211 | Class V (H-P+)<br>N = 22 | Total<br>N = 544 |
| --- | --- | --- | --- | --- |
| Age | 53.0 [47.0, 59.0] | 46.0 [39.5, 53.5] | 41.5 [37.0, 49.0] | 50.0 [42.0, 57.0] |
| Male | 199 (64.0%) | 162 (76.8%) | 14 (63.6%) | 375 (68.9%) |
| Program start before 1958 | 284 (91.3%) | 197 (93.4%) | 21 (95.5%) | 502 (92.3%) |
| Time in program, prior to<br>30-day break | 117 [53.5, 180] | 89.0 [37.0, 168] | 110 [48.3, 178] | 109 [46.0, 176] |
| Time in program, total,<br>days | 440 [148, 1530] | 180 [52.5, 697] | 552 [162, 1580] | 333 [89.0, 1110] |
| SBP, mmHg | 207 [189, 223] | 223 [210, 238] | 211 [196, 244] | 213 [194, 232] |
| Nmiss | 9 (2.9%) | 3 (1.4%) | 2 (9.1%) | 14 (2.6%) |
| DBP, mmHg | 120 [109, 134] | 137 [127, 146] | 133 [120, 141] | 128 [114, 140] |
| Nmiss | 9 (2.9%) | 3 (1.4%) | 2 (9.1%) | 14 (2.6%) |
| Weight, Kg | 69.1 [61.1, 77.9] | 68.9 [59.8, 77.0] | 76.3 [65.5, 83.3] | 69.2 [60.2, 78.1] |
| Nmiss | 197 (63.3%) | 123 (58.3%) | 16 (72.7%) | 336 (61.8%) |
| BMI, Kg/m<sup>2</sup> | 23.9 [22.0, 27.7] | 23.5 [21.4, 26.2] | 25.5 [22.0, 29.4] | 23.6 [21.5, 27.3] |
| Nmiss | 215 (69.1%) | 140 (66.4%) | 16 (72.7%) | 371 (68.2%) |
| UCl, mg/dL | 242 [117, 365] | 194 [108, 341] | 215 [102, 268] | 217 [113, 356] |
| Nmiss | 155 (49.8%) | 99 (46.9%) | 9 (40.9%) | 263 (48.3%) |
| NPN, mg/dL | 38.0 [33.5, 45.0] | 51.0 [39.0, 72.0] | 44.5 [37.0, 51.0] | 41.0 [35.0, 57.0] |
| Nmiss | 132 (42.4%) | 74 (35.1%) | 11 (50.0%) | 217 (39.9%) |
| BP Medication during first<br>4 weeks, n (%) | 23 (7.4%) | 24 (11.4%) | 1 (4.5%) | 48 (8.8%) |
| Death during first 4 | 6 (1.9%) | 17 (8.1%) | 0 (0%) | 23 (4.2%) |
| weeks, n (%) |  |  |  |  |

Median [Q1, Q3]
SBP, DBP, NPN, UCl entry: average from day -7 to day 1
Weight entry: average from day -2 to day 1

Only 48 (8.8%) of the 544 patients received antihypertensive drugs for even one day during their first four weeks on the RD; 38 were treated with a single agent; 6, with 2; and 4 with 3 agents. Medications used and number of patients taking each were: rauwolfia (22), hydralazine (12), papaverine (10), thiazide (7), guanethedine (2), pentolinium (2), mercurial diuretics (2), hexamethonium (1), spironolactone (1), ethycrynic acid (1), veratrum (1), nadolol (1). About half of the 48 patients had antihypertensive drugs discontinued during the first four weeks. During the first four weeks on the RD, 23 patients (4.2%) died.

### Blood Pressure Response and Relationship with Chloride Excretion

SBP and DBP fell continuously from entry to Week 4 (**Figure 3a**). When the BP changes for each MH class were examined separately, BP reduction relative to baseline was consistent and similar across the three classes and was already apparent by week 1 (**Figure 3b**). In addition to BP, UCl declined continuously from baseline to Week 4 across the MH classes, and the trend appears to track with the reduction of SBP/DBP (data not shown). The median weight for the cohort dropped 1.2 kg between baseline and Week 4, but notably >50% of patients were missing a recorded weight by Week 1 and >60% were missing a recorded weight during Week 4 (data not shown).

**Figure 3.**
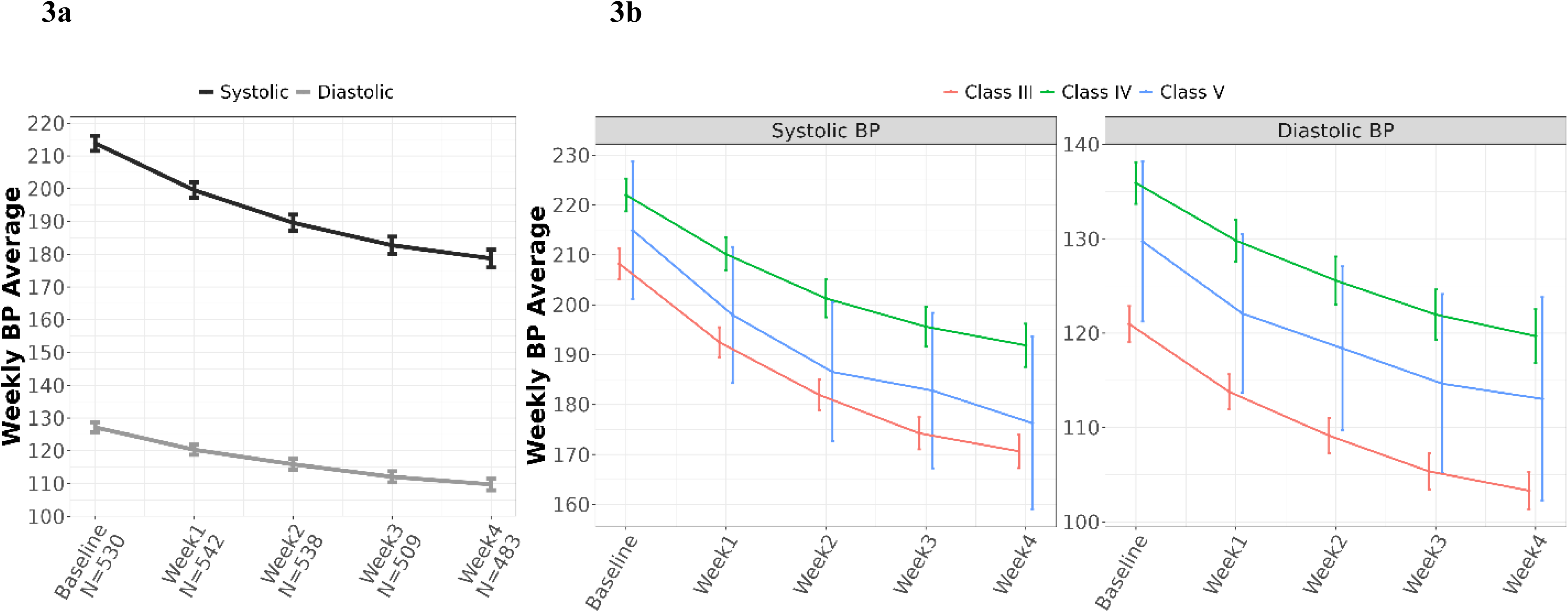
Participants’ Weekly BP from Baseline to Week 4 for the entire cohort (3a) and by MH classes (3b)

**Figure 4** shows further examination of the relationship between percent changes in SBP over 4 weeks for each RD adherence group. Those judged to have high diet adherence (ie, UCl at goal for 3 or 4 weeks) had the greatest reduction in SBP, up to an average of 5.32% reduction for each additional week in the program (beta (95% CI) = -5.06 (-5.32, -4.8) for time variable). The SBP reduction was significant over time and similar between the moderate and low adherence groups (beta:-3.79 vs. -3.27), but the level of reduction was smaller than that in the high adherence group (beta: -5.06).

**Figure 4.**
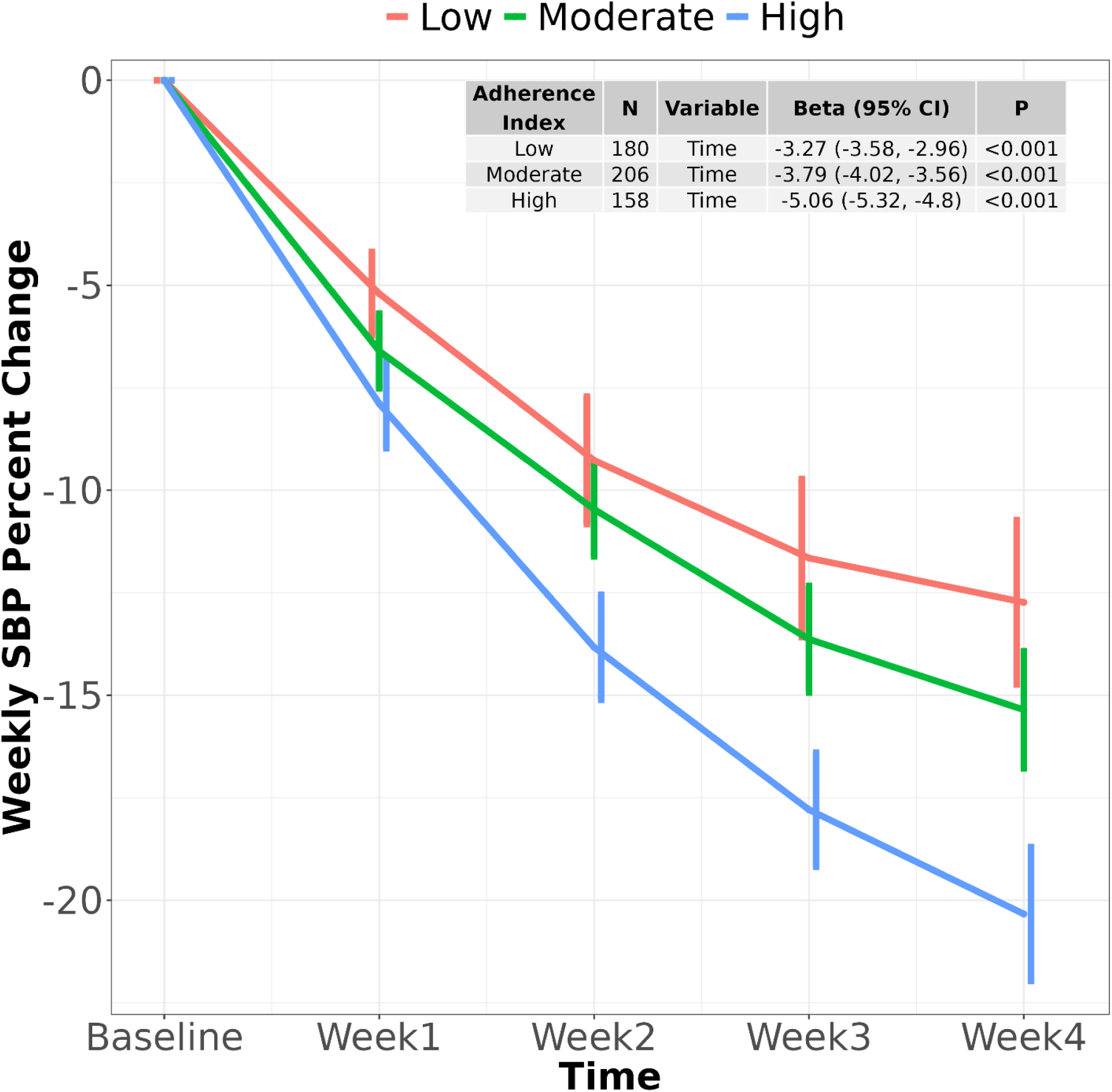
Weekly SBP changes grouped by UCl indexed adherence*. The effect of baseline SBP and time on weekly SBP change is listed by regression coefficient, beta, and its 95% confidence interval. *UCl adherence is assigned as High when meeting UCl goal of ≤42mg/dL for 3 or 4 weeks during the first 4 weeks; moderate if at goal for 1-2 weeks; or low if none of the 4 weeks at goal.

### Predictors of BP response

The relationship between baseline characteristics (age, gender, MH classification, SBP, DBP, NPN, UCl, and weight), dietary adherence (indicated by the percent of time UCl was ≤42 mg/dL, a value corresponding to a 24 hour urinary sodium excretion of approximately 250mg/day), and SBP and DBP changes from baseline to week 4 were examined using univariable and multivariable regression analyses **(Table 2).** Univariable analysis **(Table 2a**) revealed that female gender, class IV (vs. III), but not Class V, higher entry SBP, lower entry NPN, higher baseline UCl and a greater dietary adherence were all associated with a greater reduction in SBP at week 4. Only baseline DBP, UCl and dietary adherence were significantly associated with DBP changes at week 4. Class V vs class IV was not significantly associated with changes in either SBP or DBP.

**Table 2.** Univariable and multivariable linear regressions examining the change in Systolic Blood Pressures between Baseline and Week 4.

**2a Univariable:**
|  | N | SBP Change |  | DBP Change |  |
| --- | --- | --- | --- | --- | --- |
|  | 544 | Estimate (95% CI) | P | Estimate (95% CI) | P |
| Age, years | 501 | -0.13 (-0.34, 0.08) | 0.217 | 0.03 (-0.08, 0.15) | 0.602 |
| Male (vs female) | 501 | 11.57 (7.04, 16.1) | <0.001 | 1.91 (-0.66, 4.47) | 0.145 |
| Class IV (vs III) | 501 | 7.20 (2.79, 11.62) | 0.001 | 1.50 (-0.97, 3.96) | 0.233 |
| Class V (vs III) | 501 | -0.68 (-12.2, 10.9) | 0.908 | 0.93 (-5.51, 7.38) | 0.776 |
| Baseline SBP or DBP, mmHg | 501 | -0.25 (-0.33, -0.17) | <0.001 | -0.17 (-0.24, -0.11) | <0.001 |
| Baseline NPN, mg/dL | 305 | 0.18 (0.08, 0.27) | <0.001 | 0.03 (-0.02, 0.09) | 0.243 |
| Baseline UCl, mg/dL | 269 | -0.02 (-0.04, -0.01) | 0.004 | -0.01 (-0.02, 0) | 0.032 |
| UCl Compliance % | 360 | -0.31 (-0.39, -0.22) | <0.001 | -0.12 (-0.17, -0.08) | <0.001 |
| Baseline weight, Kg | 192 | -0.05 (-0.26, 0.16) | 0.646 | -0.07 (-0.19, 0.05) | 0.235 |
| Class V (vs IV) | 501 | -7.89 (-19.6, 3.83) | 0.186 | -0.56 (-7.11, 5.98) | 0.865 |

**2b Multivariable:**
|  | SBP Change |  | DBP Change |  |
| --- | --- | --- | --- | --- |
|  | N = 228 |  | N = 360 |  |
|  | Estimate (95% CI) | P | Estimate (95% CI) | P |
| Male (vs female) | 6.59 (0.03, 13.15) | 0.049 | 1.91 (-0.89, 4.71) | 0.182 |
| Class IV (vs III) | 6.23 (-0.24, 12.71) | 0.059 |  |  |
| Class V (vs III) | 0.04 (-15.7, 15.7) | 0.996 |  |  |
| Baseline SBP or<br>DBP, mmHg | -0.30 (-0.41, -0.19) | <0.001 | -0.22 (-0.29, -0.15) | <0.001 |
| Baseline NPN,<br>mg/dL | -0.03 (-0.14, 0.08) | 0.574 |  |  |
| UCI Compliance % | -0.34 (-0.46, -0.23) | <0.001 | -0.12 (-0.17, -0.08) | <0.001 |
| Class V (vs IV) | -7.49 (-24, 9.02) | 0.372 |  |  |

In multivariable analysis (**Table 2b**), women had a greater SBP reduction at week 4. In addition, higher baseline BP (either SBP or DBP) and greater dietary adherence were significantly associated with greater reductions in SBP and DBP; baseline NPN was no longer significant, suggesting that, when accounting for all variables together, baseline renal function did not significantly impact SBP reduction from sodium restriction. Based upon the multivariable analysis, an increase in dietary adherence by 10 percentage points over 4 weeks was associated with an estimated reduction of 3.4 mm Hg in SBP and 1.2 mm Hg in DBP. Furthermore, linear regression analysis revealed that a reduction in weekly UCl levels significantly predicted lower SBP during weeks 2 through 4, with the strength of this association increasing over four weeks (data not shown).

Among the 477 patients that had BP data at both baseline and week 4, the majority (85.3%, N=407) were considered “responders” (>5% BP reduction at week 4). The responders and non-responders were similar in age, baseline BP, weight and NPN, but non-responders had a higher proportion of males and a lower baseline UCl. Among the responders, 86.8% reached goal of UCl at week 4, while the comparable figure for non-responders was 55.8%. The SBP of the 70 out of 475 (14.7%) who were considered “non-responders” either increased or fell by less than 5%. Only about 16% of non-responders had high dietary adherence as indicated by UCl at goal for 3 or 4 weeks; nearly half (47%) had low adherence and 37%, moderate adherence. Sensitivity testing of multivariable regression after excluding the 48 patients that were on BP medications gave the results consistent with the main analysis (data not shown).

## Discussion

The RD offered persons with MH a chance for improved BP control at a time when there were no FDA-approved antihypertensive medications. This study describes the changes in BP for a large cohort of patients with MH, as well as patient level factors predicting a BP response to the RD. The retrospective nature of this investigation and the concurrent non-sodium dietary restrictions (i.e. fat and protein) limit precise conclusions, but the data do offer a rare opportunity to examine BP changes at levels of sodium intake well below current dietary recommendations of 1500mg/day.^15^

The BP reductions found in this study were arguably the most striking finding. By Week 4, the median SBP and DBP had fallen by 35.1 mm Hg and 16.6 mm Hg respectively. Although no modern studies compare directly with this cohort, in 1950, Watkins et al published findings from a group of 50 severely hypertensive inpatients in New York who were placed on the RD.^16^ They noted SBP/DBP changes of -29/-16 mmHg over three weeks, quite similar to the results seen in the present study, although they did note that BP in their cohort fell by -9/-3 mmHg during a 3-week ‘control’ phase prior to the initiation of the RD, suggesting that early BP changes may not be entirely attributable to the dietary intervention. In the study of Watkins et al,^16^ 8% of patients had either unchanged or a higher BP after 3-weeks on the RD. In the present study, 70 patients (14.7%) had <5% BP reduction at week 4 despite more than half of them (52.9%) adhering to the RD as indicated by UCl.

An interesting question raised by the RD data is whether the BP lowering effects are attributable to sodium restriction alone. Kempner was satisfied with the results of the diet and never investigated this aspect. However, Watkins, et al noted that adding 1g to 3g of sodium chloride to the RD reversed BP improvements in several individuals, while neither protein nor fat supplementation had an effect. Further, a meticulous study by Dole, Dahl and colleagues of 6 hypertensive patients hospitalized for 6 months on a metabolic ward and fed the RD, showed the average SBP fell from 202.7 to 161.5, which largely reversed with the addition of sodium chloride but not ammonium chloride, prompting their conclusion: “Restriction of sodium, but not of chloride appeared to be necessary for the clinical effect.”^17^ Finally, a tightly controlled study of 8 hypertensive patients by Chapman, Gibbons, and Henshel, published in 1950, demonstrated a dramatic decline in blood pressure on what they called the ‘Rice-Fruit’ diet (based on Dr. Kempner’s), with normalization of blood pressure in 2 patients and a more than 25 mmHg decline of SBP in 5 patients. They noted that the BP lowering effects of this diet were not mitigated by protein supplementation (40g) but were largely reversed by the addition of sodium (10g). Concurrent evaluation of body composition in these subjects revealed significant weight loss (1.47 kg/week over an average of approximately 4 weeks), mostly attributed to loss of extracellular fluid and fat. However, the authors also highlighted some loss of ‘active tissue’ (“presumably mostly muscle”), raising concerns about the safety and necessity of such severe protein restriction.^18^ By comparison, the DASH-Sodium trial, which evaluated three levels of dietary sodium intake (1.15 g/day, 2.3 g/day, and 3.45 g/day) using both DASH and control diets, found that lower sodium intake led to a linear reduction in BP in both diet groups. The greatest decrease in BP occurred when the DASH diet was combined with the lowest sodium intake, even though sodium restriction produced relatively less BP lowering in the DASH cohort, perhaps because the BP was already lower in the DASH than the control diet group. Taken in total, these observations raise questions about the value of fat and protein restriction in the RD.^19^

We found an inverse, linear relationship between UCl and BP, even at levels of UCl ≤42 mg/dL (i.e. estimated sodium intake ≤250mg/day), which is 6 times lower than current dietary recommendations for sodium intake (<1.5 g/day) for patients with hypertension. A rigorous meta-analysis by Fillippini et al, who examined the dose response of BP to sodium restriction in randomized trials that measured 24-hour sodium excretion (sodium range of 0.4 to 7.6g/day) provides further evidence; they also found an inverse linear relationship between BP and sodium intake, although they did not evaluate sodium intakes as low as those of the RD.^11^ These observations imply that sodium targets even lower than current guidelines might offer a potent non-pharmacologic strategy for reducing BP and mitigating vascular injury.

Multivariate analysis of the baseline characteristics of our cohort shows that female gender and a higher baseline BP best predict BP lowering on the RD. It also shows that persons with Class-III retinopathy (hemorrhage without papilledema) had a better BP lowering response to the RD than did those with Class IV retinopathy (hemorrhage with papilledema). Possibly Class-IV retinopathy reflects more advanced vascular disease or less salt sensitivity. It is interesting that higher NPN, a no-longer used marker of renal function, was inversely related to BP lowering on univariate analysis, but not on multivariate analysis, which controls for retinopathy class.

An unexpected observation from the present study regards the accrual rate of patients over the course of forty years (1942-1982). As shown in **Figure 2**, the number of patients coming to the RD increased rapidly as the existence of the program became known. However, after a peak in about 1950, there was a precipitous (near-exponential) drop in accrual until about 1958. Others have noted the “disappearance” of MH,^20^ and most^21^ assume or imply that the decreased prevalence results from the availability of effective antihypertensive medication. As is the case for some other diseases that have dramatically declined in prevalence, such as coronary heart disease^22,23^ or rheumatic fever,^24^ our data suggest that something other than pharmacotherapy is at play because the decline in patient accrual began years before effective antihypertensive medications were available. Although unlikely, we cannot rule out the possibility of a decline in patient referrals to the RD by primary care physicians.

Although rigorous methodologies were used to create and archive data from the RD program, and to conduct this analysis, there are important limitations. First, because of its retrospective nature, it is impossible to control for potential confounders. For example, this study used only observations made as part of patient care, rather than a standardized research protocol, so the timing and collection of clinical and metabolic variables were incomplete and non-uniform, which reduces statistical power and may result in measurement bias. Furthermore, it is impossible to discern from the available records which data were collected on inpatients vs. ambulatory patients, or exactly what if any, modifications were made to the RD throughout each patient’s clinical course. The paucity of patient weight data prevented examination of the possible effect of loss of extracellular fluid during salt restriction on blood pressure. Finally, although UCl is highly correlated with sodium intake, the use of spot UCl concentrations does not fully describe the relationship between sodium restriction and BP lowering.^25^

In conclusion, the RD offered patients with the dismal prognosis conferred by malignant hypertension and no other viable therapeutic options, an opportunity to lower their BP through a rigorous, albeit monotonous, dietary intervention requiring close medical supervision. The RD did effect a rapid reduction in BP, but because the RD restricted sodium, protein and fat together, it is impossible to conclude with these data that its BP lowering effects are due solely to sodium restriction. Nonetheless, these historical data suggest that patients with high blood pressure may benefit from sodium restriction many times lower than current guidelines, particularly those whose BP elevation resists available pharmacologic therapies.

## Data Availability

Data can be made available upon written request.

## Acknowledgments

SS, PJK and FAN contributed to the concept of the manuscript. SS drafted the first version of the manuscript. JOL and FAN analyzed the data. AB, DL, WM organized and provided data for analyses. PHL, SS, PJK, FAN, FCL and YJL provided critical input to analyses, interpretation of results, review and edits. PHL is the guarantor of the manuscript.

